# Stress hormones are associated with multiday seizure cycles

**DOI:** 10.1101/2025.06.11.25329394

**Authors:** Rachel E. Stirling, Jodie Naim-Feil, David B. Grayden, Wendyl J. D’Souza, Ian Gordon, Dean Freestone, Ewan Nurse, Mark J. Cook, Philippa J. Karoly

## Abstract

It is well established that most people with epilepsy experience cyclical fluctuations in seizure susceptibility. These seizure patterns have been associated multiday oscillations in cortical excitability and autonomic changes, although the mechanistic drivers of these cycles are not well understood. In this study, we measured stress hormone levels at phases of seizure cycles to investigate the autonomic system as a possible co-oscillator with multiday cycles of seizure susceptibility.

Thirteen participants with focal epilepsy were recruited for this longitudinal study. Participants reported seizures in an electronic diary for >6 months. 24 saliva samples were collected per person across two predicted high risk periods and two predicted low risk periods (“allocated risk”). Saliva samples were analysed for cortisol and dehydroepiandrosterone sulphate (DHEAS) levels. Linear mixed models were fitted to predict stress hormones with fixed effects: multiday seizure cycle (retrospective peak or trough), time of day, allocated risk, perceived stress scale score, and seizure occurrence around the saliva sample time.

Participants recorded an average of 193 (SD = 158) seizures during the study period. 312 saliva samples were collected in total. Cortisol levels were significantly higher in the epilepsy cohort compared to the expected general population. On a group level, cortisol was significantly associated with fixed effects time of day and multiday seizure cycle, with cortisol levels heightened at multiday cycle peaks compared to troughs, particularly evident in the morning saliva samples.

These results provide new insights into a possible mechanistic driver or co-oscillator of multiday seizure cycles in people with epilepsy, demonstrating that cortisol levels are higher at the peak of multiday cycles irrespective of seizure occurrence. The novel methodology presented in this work may be used to explore interactions between other biomolecules of interest and multiday seizure cycles.

## Introduction

Epilepsy is a prevalent and debilitating neurological disorder characterised by recurrent unprovoked seizures, with stress and stress hormones identified as a major component of epilepsy. Understanding the dynamic relationship between stress and seizure activity is challenging due to complex and bi-directional interactions between them^1^. Stress is frequently self-reported as a major trigger for epileptic seizures^2^, with seizure diary studies linking stressful life events with increased seizure frequency^3–6^. Acute increases in cortisol can precipitate epileptic seizures^7^, while chronic stress and sustained cortisol elevation increase seizure frequency^8^. Seizures can also act as stressors, triggering autonomic nervous system changes^8^ and activating the hypothalamic-pituitary-adrenal axis^9,10^, leading to elevated cortisol levels^7,11^. Compared to healthy controls, basal cortisol levels are often reported to be higher in individuals with epilepsy^12–14^. However, findings remain somewhat mixed, as other studies have reported reduced cortisol levels^15,16^ or no difference^17,18^. A deeper understanding of the long-term dynamic interplay between stress markers and seizures may enable a unifying explanation for these observations.

Epileptic seizures were long considered unpredictable events with no discernible pattern. However, recent advances in long-term intracranial EEG monitoring have revealed that both seizures and interictal epileptiform activity follow individual-specific daily and multiday (>2 day cycle) cycle patterns^19^, occurring in about 90% of people with epilepsy^20,21^. The discovery of rhythmic fluctuations in seizure susceptibility has pioneered a new direction in epilepsy research, with cycle-based seizure risk models consistently outperforming other seizure prediction methods^22–25^. Notably, seizure cycles can be estimated non-invasively using self-reported seizure diaries^22,26^, demonstrating patterns comparable to those observed in intracranial EEG recordings^22,23,27,28^. Moreover, multiday cycles extend beyond neural activity, with similar rhythmic patterns observed in other physiological markers, such as heart rate^29,30^, suggesting that these fluctuations may be driven by intrinsic biological processes governing multiple interconnected physiological systems, similar to circadian processes. Thus, to understand whether fluctuations in stress markers, like cortisol, align with periods of heightened seizure likelihood, it is critical to examine stress longitudinally across these individual-specific cycles. This could unlock new avenues for targeted interventions aimed at mitigating stress during high-risk seizure periods.

Cortisol levels are regulated by the body’s natural circadian rhythm, which in turn plays a significant role in maintaining metabolic homeostasis, modulating hormone responses, and influencing the sleep-wake cycle^31,32^. One study found that circadian fluctuations in cortisol secretion modulate the daily timing of interictal epileptiform activity^33^. At slower timescales, cortisol levels appear to follow multiday rhythms, particularly seasonal and menstrual cycles in the general population^13,34–36^. However, a very limited number of studies have explored whether these rhythms are dysregulated in epilepsy^13^ or stressed-invoked states^37^. To the authors’ knowledge, no previous studies have explored how these stress markers fluctuate over multiday seizure cycles. This gap in the research exists because most existing cortisol studies are conducted over shorter timeframes and lack sufficient data to detect long-term trends (over weeks to months). Additionally, it is often infeasible or impossible to continuously monitor biomolecules (such as cortisol), over extended periods. However, novel approaches for the extraction and projection of individual-specific seizure cycles now provide a methodology capable of tracking how an individual’s stress hormones fluctuate over different phases (i.e., the peak versus the trough) of their multiday cycle, without requiring long-term, continuous monitoring. This approach may reveal previously difficult-to-obtain insights into the longer-term stress-seizure dynamics in epilepsy.

The present study piloted a longitudinal design using cycle-based modelling to examine the interaction between stress hormones (cortisol and dehydroepiandrosterone sulfate (DHEAS)), perceived stress levels (self-perceived stress score) and individual-specific multiday seizure cycles. The study aimed to 1) identify whether stress markers such as cortisol, DHEAS and self-perceived stress fluctuate over multiday seizure cycles, and 2) examine whether additional factors, such as time of day and seizure occurrence, impact the relationship between stress and seizure cycles. This novel study is the first investigation of how stress fluctuates at multiday timescales in people with epilepsy.

## Methods

Eighteen adults with a clinical diagnosis of focal epilepsy were recruited from the neurology clinic at St Vincent’s Hospital, Melbourne (Australia) and from Seer Medical (Melbourne, Australia). Some of these participants were concurrently enrolled in a separate study investigating the effectiveness of scheduling diagnostic EEG sessions during high seizure risk periods^38^.

To be eligible for inclusion, participants were required to have reported at least 10 seizure events in an electronic seizure diary (Seer app, Seer Medical) over at least 6 months prior to the start of salivary sampling. Of the 18 participants initially recruited, five did not meet the inclusion criteria or were lost to follow-up due to the demands of the study protocol, including its complexity and time commitment. Clinical and demographic characteristics of participants who completed the full study protocol (n=13) are provided in Table 1.

**Table 1.**
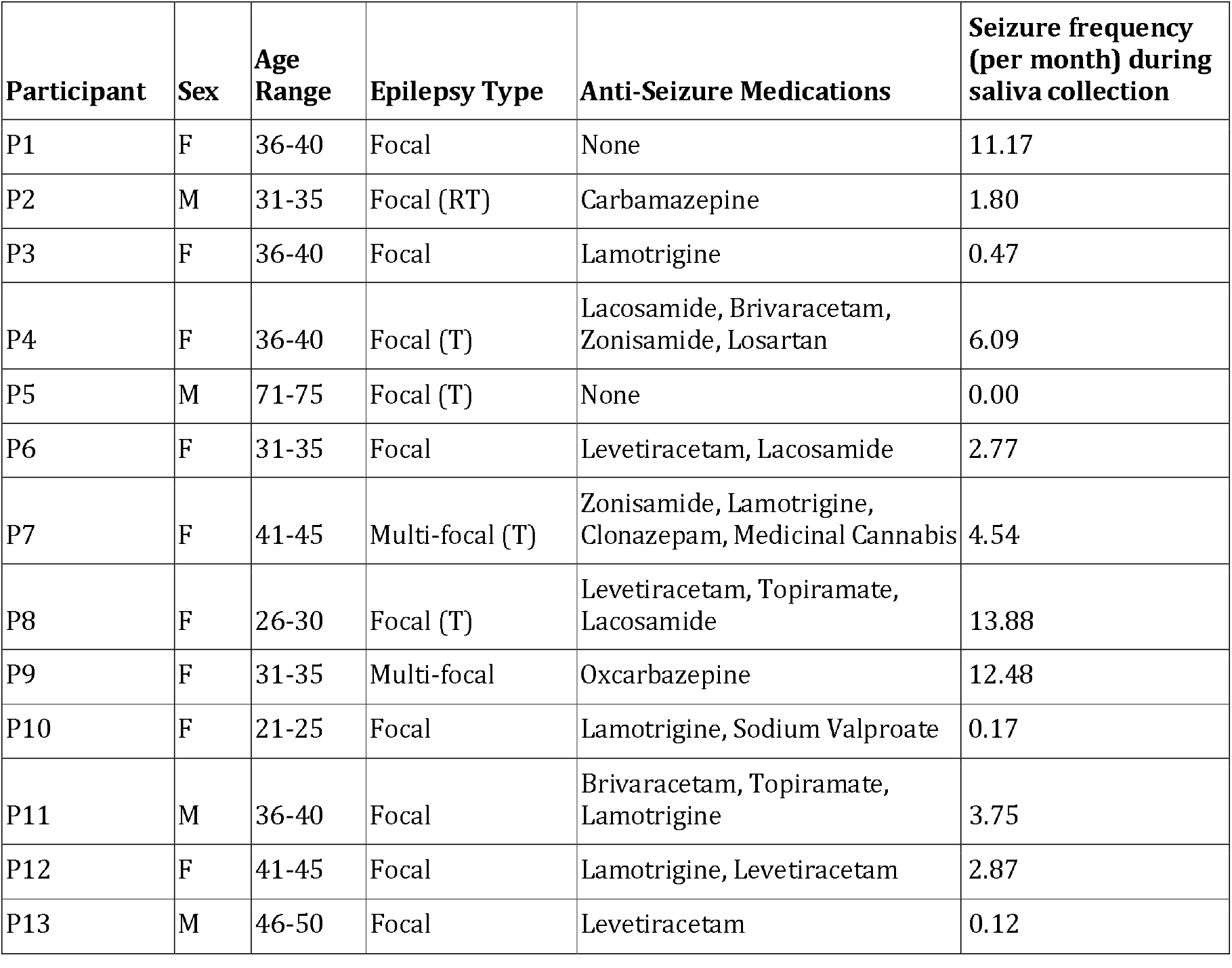
Demographic and clinical characteristics of study participants. T=Temporal, RT=Right Temporal

| Participant | Sex | Age Range | Epilepsy Type | Anti-Seizure Medications | Seizure frequency (per month) during saliva collection |
| --- | --- | --- | --- | --- | --- |
| P1 | F | 36-40 | Focal | None | 11.17 |
| P2 | M | 31-35 | Focal (RT) | Carbamazepine | 1.80 |
| P3 | F | 36-40 | Focal | Lamotrigine | 0.47 |
| P4 | F | 36-40 | Focal (T) | Lacosamide, Brivaracetam, Zonisamide, Losartan | 6.09 |
| P5 | M | 71-75 | Focal (T) | None | 0.00 |
| P6 | F | 31-35 | Focal | Levetiracetam, Lacosamide | 2.77 |
| P7 | F | 41-45 | Multi-focal (T) | Zonisamide, Lamotrigine, Clonazepam, Medicinal Cannabis | 4.54 |
| P8 | F | 26-30 | Focal (T) | Levetiracetam, Topiramate, Lacosamide | 13.88 |
| P9 | F | 31-35 | Multi-focal | Oxcarbazepine | 12.48 |
| P10 | F | 21-25 | Focal | Lamotrigine, Sodium Valproate | 0.17 |
| P11 | M | 36-40 | Focal | Brivaracetam, Topiramate, Lamotrigine | 3.75 |
| P12 | F | 41-45 | Focal | Lamotrigine, Levetiracetam | 2.87 |
| P13 | M | 46-50 | Focal | Levetiracetam | 0.12 |

Saliva samples were collected at four predetermined time periods for each participant throughout the study, to investigate changes in stress hormone levels over time. Each saliva sampling collection period lasted for three days and involved six saliva collections; thus, each participant provided 24 saliva samples over the study duration. Two stress-related hormones were measured in the saliva samples: cortisol and dehydroepiandrosterone sulfate (DHEAS). Salivary sampling was chosen for its non-invasive and convenient nature and is considered the gold standard for assessing free cortisol, which reflects the biologically active, unbound hormone fraction^39^. Ethical approval for this study was granted by St Vincent’s Hospital Human Research Ethics Committee (HREC 009.19). All participants provided informed consent and were made aware that participation was voluntary, with the option to withdraw at any time.

### Study design

A schematic of the study design and sampling protocol is shown in Figure 1. To schedule the four saliva sample collection periods, a cycle-based seizure forecasting algorithm was used to identify the highest-risk or lowest-risk days for seizure occurrence within a three-month window, based on each participant’s unique seizure cycles (see *Allocated Risk*). Each saliva sample collection period involved saliva collection on three consecutive days: the day before the identified high or low risk day, and the two following days. This procedure was repeated across four time points (two during high-risk periods and two during low-risk periods), resulting in four three-day sampling rounds. Participants provided six saliva samples per round (morning and evening on each of the three days), totalling 24 samples across the study. All participants were blinded to their risk allocation.

**Figure 1.**
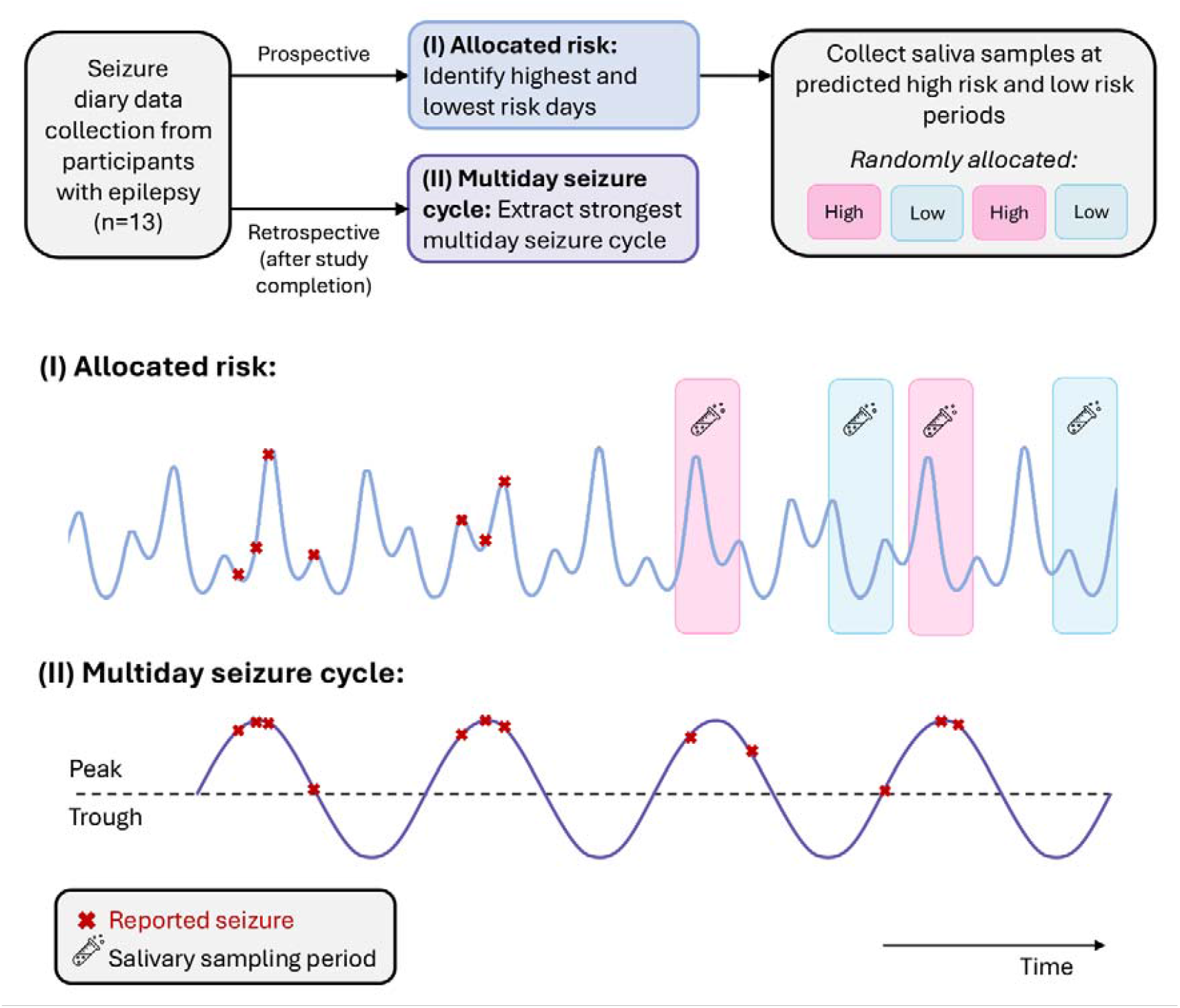
Study protocol schematic. Prior to salivary sampling, participants reported at least 10 seizures in an electronic seizure diary over a minimum of 6 months. Using this data, the highest and lowest risk days (two each) were identified in the next three months using a seizure risk forecasting algorithm (see *Allocated Risk*). Participants were randomly assigned to salivary sampling during either a high risk or low risk period. Saliva samples were collected during two high risk periods and two low risk periods, each sampling round conducted over 3 days. Participants continued to use their seizure diary during the sampling phase of the study, with forecasts updated between sampling periods. At study conclusion, the seizure diary was used to extract the strongest multiday seizure cycle (retrospectively). **(I)** An example seizure risk forecast (generated using seizures reported before saliva sampling), which identified two predicted high risk days (red shading) and two low risk days (green shading), during which the saliva sampling was completed. Note that seizures (red crosses) are plotted only before saliva sampling occurred to demonstrate that only seizures reported before saliva sampling were used by the forecast to predict high and low risk sampling periods. **(II)** A schematic representation of a multiday seizure cycle retrospectively fitted to the whole duration of the seizure diary (i.e., seizures reported during the whole study: before and during saliva sampling).

**Figure 2.**
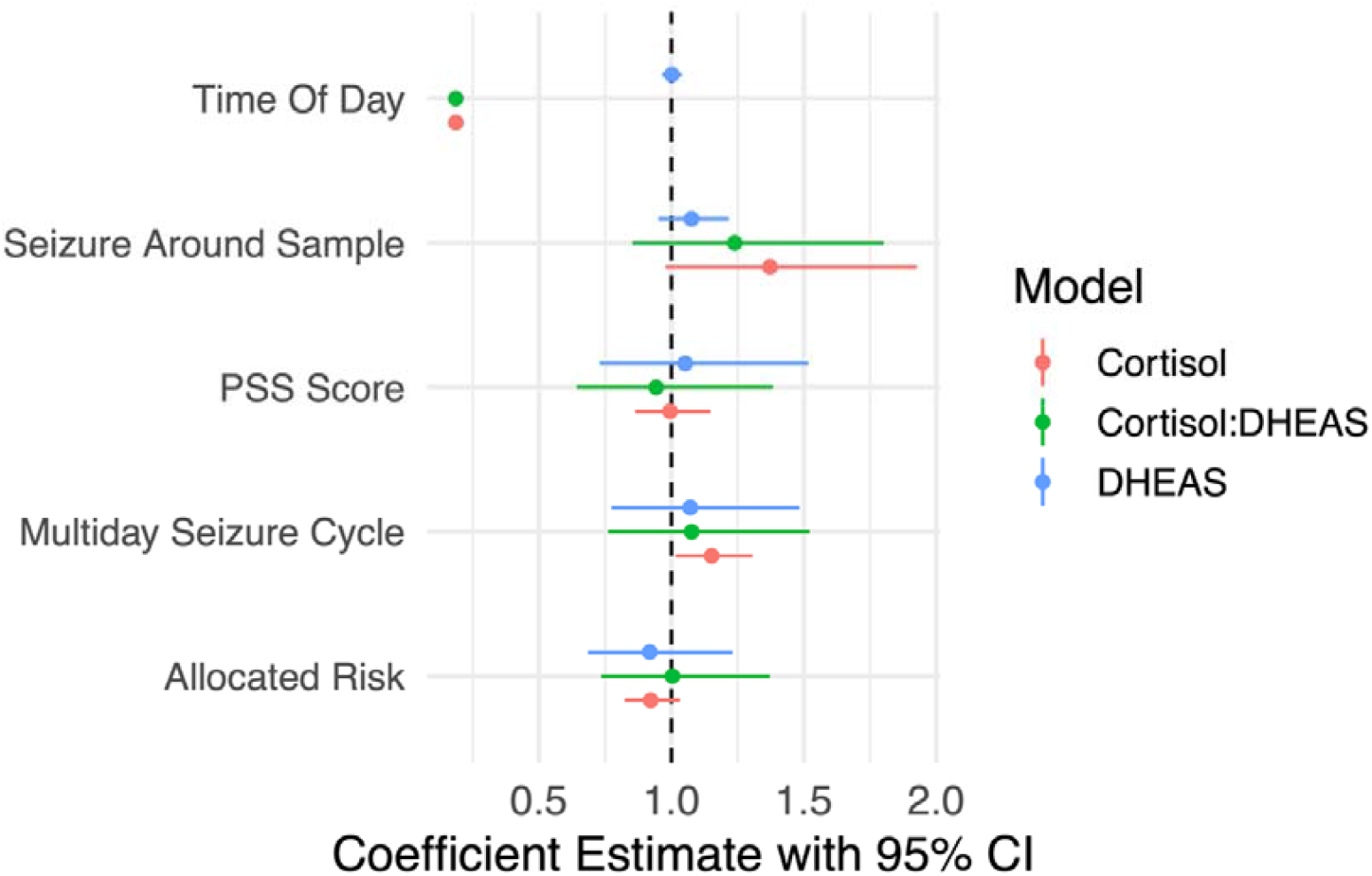
Coefficient estimates and 95% confidence intervals (CIs) for fixed effects in the linear mixed models (Equation 1). Three models were evaluated and plotted together: Cortisol concentration (red), Cortisol:DHEAS ratio (green) and DHEAS concentration (blue). Coefficient estimates and confidence intervals were exponentiated, giving a ratio measure of effect on the response variables.

During each three-day sampling round, saliva samples were collected at 8 AM and 8 PM on each day. Participants were instructed to avoid brushing their teeth or consuming caffeine, food, or alcohol before the morning saliva sample and within two hours prior to the evening sample. Saliva samples were analysed for cortisol and DHEAS levels. Cortisol was measured in all six saliva samples per round to capture known diurnal fluctuations in cortisol, while DHEAS was measured only in the Day 1 morning (8 AM) sample, as it is more stable due to its longer half-life and minimal diurnal variation^40^.

To assess perceived stress levels, participants also completed the validated 10-item Perceived Stress Scale (PSS)^41^ at the start of each sampling round. The PSS was used to determine whether perceived stress levels were associated with stress hormone concentrations over repeated measures^42^, exploring the potential of the PSS as a marker of physiological stress, an association previously reported in other populations^43^ but not yet investigated in people with epilepsy. The scoring system for the PSS is reported in Supplementary Table 1.

### Allocated risk

Risk period allocation followed the methodology outlined in previous work^44^. For each participant, a cycle-based forecasting algorithm was trained on the individual’s seizure diary data reported prior to the initiation of saliva sampling. Seizure diary data were processed and produced daily seizure likelihood values for 3 months into the future.

Seizure cycles were extracted from the seizure diary using the method presented in previous work^38,45^. Synchronization Index (Equation 1) values were calculated for each possible cycle period between 3 and 70 days in increments of 12 hours. The Synchronization Index is

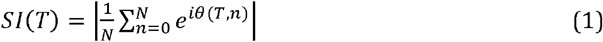

where *N* is the total number of reported seizures, *n* is the seizure number in the sequence of reported seizures, *θ* (*T, n*) is the phase of the *n*^th^ seizure relative to the cycle period *T*, and *i* is the imaginary number. The forecasting algorithm selected the two strongest cycles (based on their Synchronisation Index values, requiring SI≥0.3) between 3 and 70 days to forecast seizure risk. These parameters were chosen to align with a separate, ongoing study investigating the effectiveness of scheduling diagnostic EEG sessions during high seizure risk periods^38^ in which some participants were concurrently enrolled.

The two strongest seizure cycles were projected forward in time (beyond the last seizure diary entry) using a Von Mises risk distribution model, which assumes that seizure cycles are sinusoidal in nature and have a fixed period. The von Mises risk distribution model is derived for each cycle period by mapping historic seizure times to cycle phases to produce a phase-dependent seizure likelihood distribution. Fixed seizure cycles were extrapolated beyond the seizure diary dates, and the von Mises risk distribution model was used to estimate daily seizure likelihoods. Daily seizure likelihoods for the two strongest cycles were combined using a log odds model to produce the final daily seizure likelihood forecast for 3 months into the future.

Salivary sampling round dates were chosen by selecting two highest and two lowest seizure likelihood days in the next 3 months, respectively, provided that consecutive sampling rounds were separated by at least three days. Each sampling round lasted for three days and was conducted around the highest or lowest risk date (day before, day of, day after). The order in which these risk states were completed was random, based on participant availability and convenience.

### Multiday seizure cycle

After the study was completed and all salivary samples were collected, the strongest seizure cycle was *retrospectively* modelled from the complete seizure diary, using seizure events reported prior to, between and during salivary sampling periods. Incorporating all reported seizure events strengthens the accuracy of the multiday seizure cycle model, as the prospective *Allocated Risk* forecast relies on the assumption that biological rhythms can be projected using fixed period models, which does not appear to hold true in real-world biological systems.

To determine the strongest seizure cycle, the Synchronization Index was calculated on all possible cycle periods between 4 and 200 days, in increments of 12 hours. The strongest seizure cycle (i.e. highest SI value) was fitted to the seizure diary using a von Mises risk distribution model, whereby cycle peaks represent highest seizure risk days and troughs represent lowest seizure risk days. Once the strongest multiday cycle was modelled, salivary sampling periods were categorized as occurring during either during a cycle peak or a cycle trough. This classification was based on the cycle phase on the second (middle) salivary sampling day, with peaks and troughs defined as days landing above or below the 50th-percentile of all cycle likelihoods, retrospectively (Figure 1). The binary classification (peak, trough) of multiday seizure cycle phase was ideal for the smaller sample size used in the current work but could be expanded to incorporate more phases in a larger cohort.

### Data Analysis

#### Linear mixed models

Linear mixed models (with Nelder-Mead optimisers) were fitted to predict log-transformed concentrations of the biomolecules of interest (Equation 2): cortisol concentration (nmol/L), DHEAS concentration (nmol/L) and the derived cortisol:DHEAS ratio. Each model included the following variables as fixed effects: time of day (morning vs evening), allocated risk period (high vs low), seizure around sample (whether or not a seizure occurred within ±1 hour of the saliva sample), multiday seizure cycle (peak vs trough) and Perceived Stress Scale score. The model included saliva sampling collection period (i.e., 1, 2, 3, or 4) as a random effect, nested under each participant,

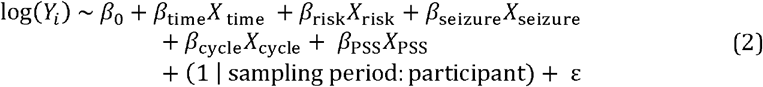

where *Y*_*i*_ denotes the concentration of the biomolecule of interest, *β*_0_ is the fixed intercept, ε is the error, *β*_*i*_ are the fixed effect coefficients and *X*_*i*_ are the fixed effect variables representing:

› **Time of day (*X*_time_):** whether the saliva sample was provided in the morning (0) or night (1).
› **Allocated risk (*X*_risk_):** whether the saliva sample was given during an allocated high (1) or low (0) risk period. This date was forecasted up to 3 months before the sampling period. This is similar to intention-to-treat analysis in clinical trials and evaluates the real-world effectiveness of measuring biomolecules at assigned risk states.
› **Seizure around sample (*X*_seizure_):** whether (1) or not (0) a seizure was reported within 1 hour before or after the saliva sample was provided, as pre- and post-ictal changes in cortisol have previously been reported^17,46^.
› **Multiday seizure cycle (*X*_cycle_):** whether the salivary sampling period was conducted at a multiday seizure cycle peak (1) or trough (0), retrospectively.
› **Perceived stress scale (PSS) score (*X*_PSS_)**: total PSS score, where higher values indicate higher perceived stress levels (Supplementary Table 1). This fixed effect was scaled per 10 units (i.e., divided by 10) to ensure confidence intervals were comparable to other variables.

Cortisol, DHEAS and Cortisol:DHEAS ratio were log transformed prior to model fitting to reduce the effect of skewed data.

Models were fitted using the lme4 package in R, with random intercepts specified for each participant and saliva sampling collection period to account for repeated measures. Fixed effects significance was assessed using ANOVA with Kenward-Roger correction for adjusting degrees of freedom.

#### Other analyses

Sub-analyses were conducted to further understand the relationships observed in the linear mixed models. Significance of slopes in Figure 3A were tested using a linear regression analysis. To explore factors contributing to different slopes, we constructed linear models predicting the slope of cortisol change across multiday cycles, stratified by time of day (morning vs evening samples). Stepwise regression using backward selection was performed to iteratively remove non-informative predictors based on Akaike Information Criterion. Separate models were fitted for morning and evening samples using predictors: intra-individual variability in cortisol and DHEAS levels, multiday cycle strength (i.e., Synchronization Index) and circadian seizure chronotype (mean phase of seizure timing on circadian clock, where 1= morning phase, i.e., 12AM-12PM, and 0 = afternoon phase, i.e., 12PM-12AM) (Supplementary Figure 1).

**Figure 3.**
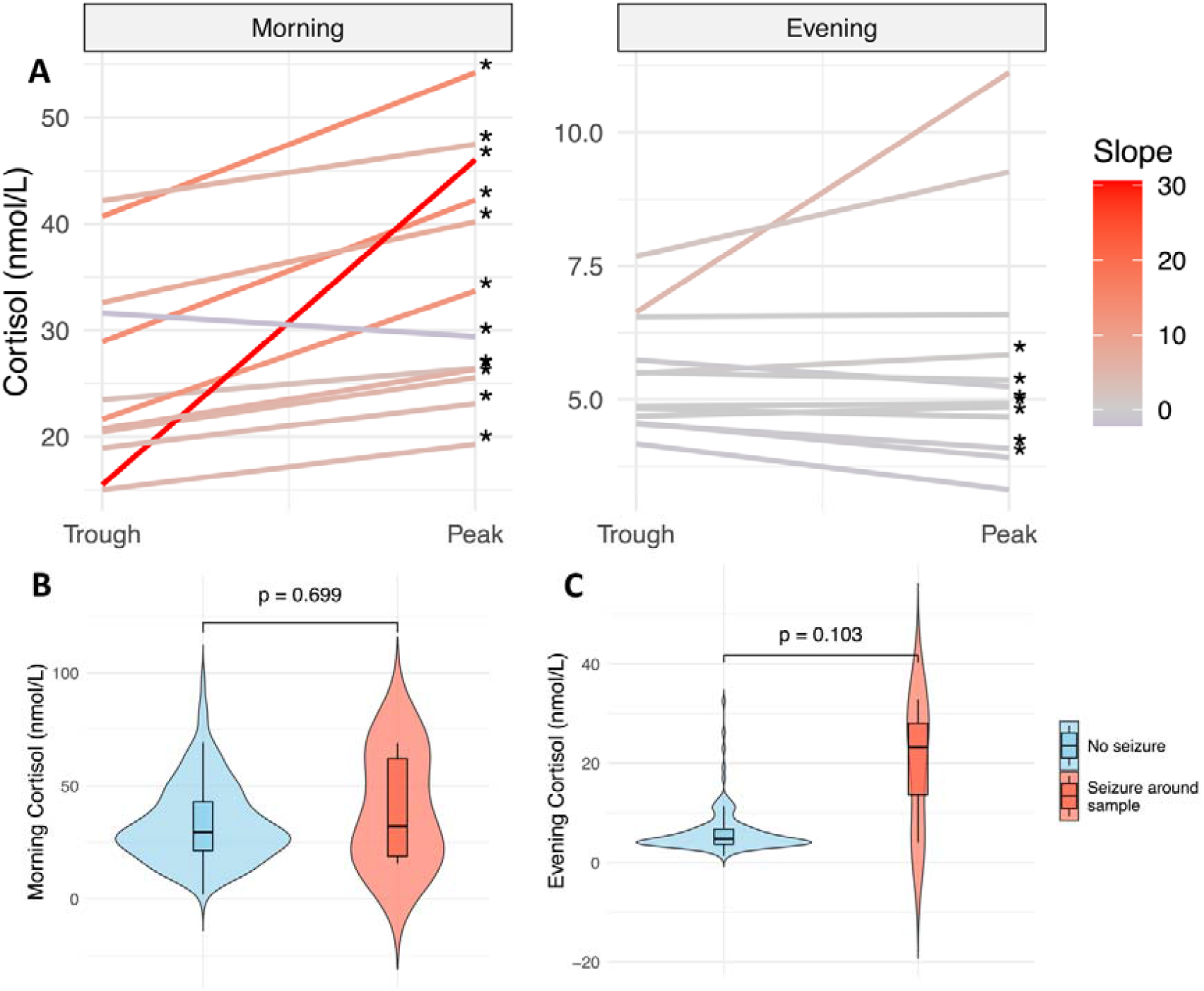
Morning and evening cortisol concentrations (nmol/L) at multiday cycle peaks and troughs, and in response to seizure timing. (A) Individual linear regression models for cortisol concentrations recorded around cycle troughs (left hand side of x axes) compared to cycle peaks (right hand side of x axes), where the colour of the fitted line represents the strength of the slope. Significance (*, p < 0.05) of these lines was tested using linear regression (t-test) analyses. (B-C) Morning and evening cortisol concentrations recorded around seizure-free times (blue) and around seizure times (red), where seizure times are defined as a seizure reported within 1 hour either side of the time the salivary sample was provided.

Median values of morning and evening hormone concentrations (Figure 3B-C) and PSS scores (Figure 4) across various parameters were compared using the non-parametric Wilcoxon rank-sum test (p-values reported). Differences in the distributions of stress hormone levels across the population at multiday cycle phase and time of day (Supplementary Figures 2-4) were also tested, using the Kolmogorov-Smirnov test and p-values were reported. The Pearson’s correlation test was used to measure the correlation between items on the PSS scale and hormone levels (Supplementary Figure 5) and seizure frequency and hormone levels.

**Figure 4.**
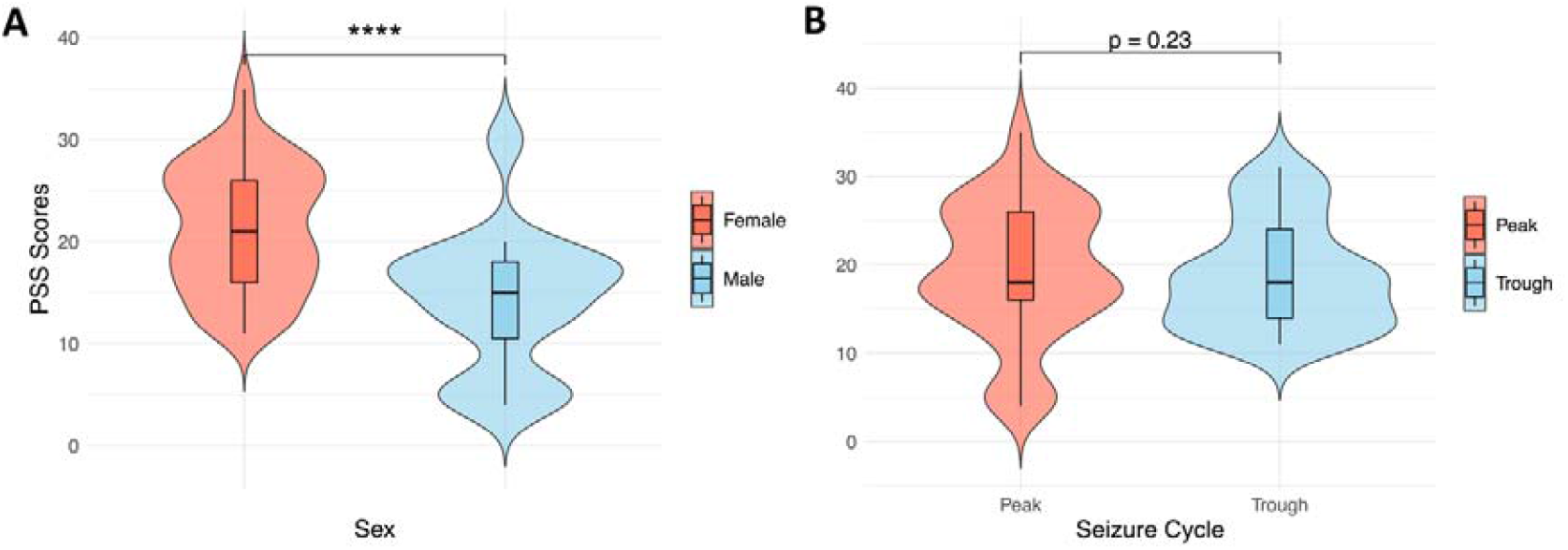
The relationship between total Perceived Stress Scale (PSS-10 questionnaire) scores and sex (A) and multiday seizure cycles (B) across the epilepsy cohort. (A) Red box plots represent PSS scores reported by females, while blue box plots represent PSS scores reported by males. (B) Red box plots represent PSS scores recorded during peaks in multiday seizure cycles, and blue box plots represent PSS scores recorded during troughs.

All statistical analyses were conducted in R (version 4.4.0).

## Results

Thirteen (n=13) participants with epilepsy were included in the analysis (see Table 1 for demographic and clinical characteristics). Participants recorded a mean of 146 (standard deviation (SD) = 158) seizures over 29.3 months (SD = 31.8) prior to beginning saliva sampling, and a mean of 47 (SD = 62) seizures between their first saliva collection and the end of the study (duration mean ± SD: 11.8 ± 8.0 months). All participants provided 24 saliva samples (12 morning and 12 night) throughout the study. Participants’ mean morning cortisol, evening cortisol and DHEAS concentrations were 33.7nmol/L (SD = 11.5nmol/L), 6.4 nmol/L (SD = 2.7nmol/L) and 14.9nmol/L (SD = 10.3nmol/L), respectively.

Linear mixed models (Equation 1) were fitted to predict log-transformed Cortisol concentration, Cortisol:DHEAS ratio and DHEAS concentration using the fixed effects of time of day, multiday seizure cycle, allocated risk, seizure around sample and PSS score, with saliva sampling collection period specified as a random effect. Coefficient estimates, confidence intervals and p-value results for all models are shown in Figure 2 and Table 2.

**Table 2.**
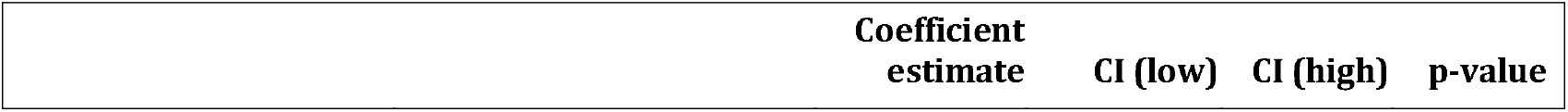
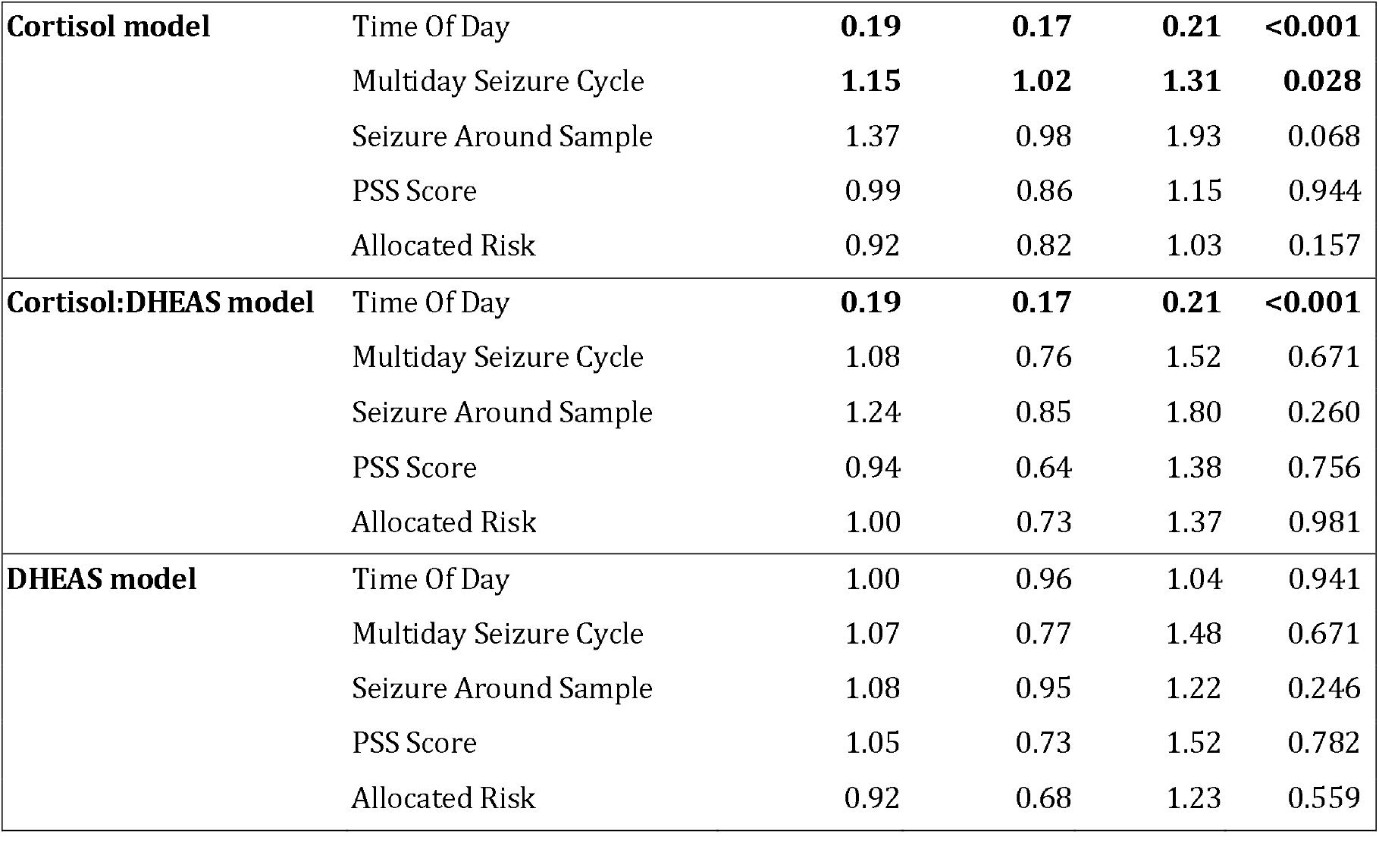
Coefficient estimates, confidence interval (CI) and p-values values for variables in the Cortisol, Cortisol:DHEAS and DHEAS linear mixed models. Significant (p < 0.05) values are bolded. Coefficient estimates and confidence intervals were exponentiated, giving a ratio measure of effect on the response variables.

|  |  |  |  |  |  |
| --- | --- | --- | --- | --- | --- |
| **Cortisol model** | Time Of Day | **0.19** | **0.17** | **0.21** | **<0.001** |
|  | Multiday Seizure Cycle | **1.15** | **1.02** | **1.31** | **0.028** |
|  | Seizure Around Sample | 1.37 | 0.98 | 1.93 | 0.068 |
|  | PSS Score | 0.99 | 0.86 | 1.15 | 0.944 |
|  | Allocated Risk | 0.92 | 0.82 | 1.03 | 0.157 |
| **Cortisol:DHEAS model** | Time Of Day | **0.19** | **0.17** | **0.21** | **<0.001** |
|  | Multiday Seizure Cycle | 1.08 | 0.76 | 1.52 | 0.671 |
|  | Seizure Around Sample | 1.24 | 0.85 | 1.80 | 0.260 |
|  | PSS Score | 0.94 | 0.64 | 1.38 | 0.756 |
|  | Allocated Risk | 1.00 | 0.73 | 1.37 | 0.981 |
| **DHEAS model** | Time Of Day | 1.00 | 0.96 | 1.04 | 0.941 |
|  | Multiday Seizure Cycle | 1.07 | 0.77 | 1.48 | 0.671 |
|  | Seizure Around Sample | 1.08 | 0.95 | 1.22 | 0.246 |
|  | PSS Score | 1.05 | 0.73 | 1.52 | 0.782 |
|  | Allocated Risk | 0.92 | 0.68 | 1.23 | 0.559 |

For Cortisol, the model explained 76% of the variance ( ) and 71% by fixed effects alone ( ). The effects of time of day and multiday seizure cycle were statistically significant (p < 0.001 and p = 0.028, respectively). These effects were also significant at the cohort level (Supplementary Figures 2 and 3). While seizure around sample was positively correlated with cortisol concentration, this relationship was not statistically significant (p = 0.068), likely due to the variability in inter-participant stress responses to seizures.

For Cortisol:DHEAS ratio, the model explained 84% of the variance ( ) and 45% by fixed effects alone ( ). Like Cortisol, the effect of time of day was significant (p < 0.001), which is entirely explained by the proportional relationship between Cortisol and Cortisol:DHEAS ratio.

For DHEAS, the model’s explanatory power was substantial (, but the contribution from fixed effects was negligible ( ) and none of the variables had a statistically significant effect (p > 0.05).

Given the strong effect of time of day observed in the Cortisol model, cortisol concentrations at multiday cycle peaks and troughs were analyzed separately for morning and evening samples (Figure 3A). One participant (P5) was excluded from the analysis as their samples were collected only at retrospective peaks. In morning saliva samples, cortisol levels were significantly elevated at multiday cycle peaks compared to troughs in 11 out of 12 participants. In evening saliva samples, cortisol was significantly elevated at cycle peaks compared to troughs in 3 out of 12 participants but was significantly reduced at peaks in 4 out of 12 participants.

To better understand the factors influencing individual slopes (Figure 3A), we examined predictor effects separately for morning and evening samples (Supplementary Figure 1 and Supplementary Table 2). In morning samples, multiday cycle strength was identified as a significant positive predictor of slope (p = 0.047), highlighting a “dose-dependent” relationship whereby stronger multiday rhythms correspond to steeper slopes (i.e., higher cortisol levels at peaks relative to troughs). Intra-individual variance in DHEAS was also a significant positive predictor of slope in both morning and evening samples, whereas higher intra-individual variance in cortisol had a negative association with slope in evening samples but no significant association with morning slopes. Circadian seizure chronotype, i.e. whether seizures tended to occur in the morning or evening, did not appear to influence slope steepness in this cohort.

Across the cohort, participants had marginally higher DHEAS levels compared to the expected general population (Supplementary Figure 4), which was significant at cycle peaks (p = 0.018), not troughs. In contrast, participants had substantially higher cortisol levels compared to the expected general population (p < 0.001), regardless of whether the sample was taken at a multiday seizure cycle peak or trough (Supplementary Figure 2) or in the morning or evening (Supplementary Figure 3). These observations may, in part, be attributed to the presence of seizures in people with epilepsy, as evening cortisol levels did appear to be higher around times of seizures (Figure 3C), although this trend was only observed in evening saliva samples and was not significant (p = 0.103).

Seizure frequency (in seizures per month, including seizures reported between participants’ first saliva collections and the end of the study) was also compared to stress hormone concentrations. No significant associations (p>0.05) were found between seizure frequency and the mean or variance of participants’ stress hormone (DHEAS, morning cortisol and evening cortisol) concentrations throughout the study.

The average Perceived Stress Scale (PSS) score across the epilepsy cohort was 19 (SD = 7, Males=14.1±6.6, Females=21.3±6.1) (Supplementary Figure 5), which is higher than the general population (12.1 for males and 13.7 for females)^47^. Total PSS scores were significantly higher in females compared to males (Figure 4A), consistent with previous findings^41,47^. No significant relationship was observed between the peaks and troughs of multiday seizure cycles and total PSS scores (Figure 4B), indicating that perceived stress does not vary systematically with multiday seizure cycles.

Additionally, a sub-analysis of items on PSS questionnaire items and stress hormone concentrations revealed that item 9 (*“In the last month, how often have you been angered because of things that were outside of your control?”*) was the only item to be positively correlated with DHEAS (R value = 0.304, p = 0.028) and cortisol levels (R value = 0.124, p = 0.028) (Supplementary Figure 6).

## Discussion

This study investigated stress hormones and multiday seizure cycles in a longitudinal pilot study of 13 individuals with epilepsy. The findings provide a pivotal step towards identifying the dynamic interplay of stress and seizures over long timescales, which is fundamental to understanding the association of stress with seizure susceptibility, in addition to seizure occurrence, and may also shed light on mechanisms of multiday rhythms in epilepsy.

Across the cohort, cortisol levels were significantly higher during multiday cycle peaks (increased seizure susceptibility) compared to cycle troughs (decreased susceptibility), an effect which was more pronounced in individuals with stronger cyclical trends in seizure timing. This increased cortisol may be partly induced by seizure occurrence, which is supported by previous studies^7,11^. However, the current study found acute seizure-related changes in cortisol were not significant, despite a trend for increased evening cortisol around seizure occurrence, nor was seizure frequency correlated with overall cortisol. Therefore, it is possible that, at least in some individuals with epilepsy, the increase in stress hormones precedes (and drives) cyclic patterns of seizure susceptibility^7,11^.

Previous studies that investigated stress hormones in epilepsy primarily assessed either baseline cortisol and/or DHEAS levels in people with epilepsy, compared to healthy controls, or investigated short-term changes in cortisol before and after seizures^7^. Most studies show an acute, post-ictal increase in cortisol, suggesting seizures act as a stressor, although this effect has primarily been demonstrated from serum cortisol. Notably a pivotal study by van Campen et al^33^ assessed the temporal correlation between salivary (circulating) cortisol and rate of epileptiform discharges, revealing their relationship over circadian timescales, particularly in people who reported stress-sensitive seizures. The current study expands this work by considering the multiday dynamics between cortisol and seizures.

Very few previous studies have assessed long-term, individual changes in cortisol. One early study assessed longitudinal serum cortisol at three-month intervals in 45 patients with newly diagnosed epilepsy and found pronounced variability in group average cortisol levels over the two-year study, despite consistent anti-seizure medication dose^15^. Unfortunately, the aforementioned study did not report on individual trends; group level variability may have been driven by intra-individual multiday fluctuations in cortisol. To our knowledge, no other studies have tracked cortisol dynamics in people with epilepsy over multiple days. However, a longitudinal study tracked daily self-reported stress for over a year in 71 people with epilepsy^6^ and found stress levels were positively associated with seizure risk. Other studies of physiological signals linked to the stress response (heart rate, electrodermal activation, skin temperature) also identified intrinsic multiday rhythms linked to seizure cycles^29,30^. These previous studies, alongside the presented results, highlight the importance of considering dynamic interactions between stress and seizures.

In addition to disruption of cortisol associated with seizure occurrence or seizure cycles, people with epilepsy may exhibit higher baseline levels of stress hormones compared to control groups^12–14^. Furthermore, people with epilepsy show increased self-reported stress and anxiety compared to the general population^48^. The current cohort showed higher cortisol levels compared to the distribution of the expected general population, regardless of when cortisol was measured (i.e., at the peak or trough of their seizure cycle). These findings support previous studies proposing epilepsy as a model for chronic stress, with further sensitization induced by seizures^7–11^. Perceived stress was also higher in the epilepsy cohort than the general population, and higher in females than males, consistent with previous reports^41^. Interestingly perceived stress was not linked to seizure cycles or stress hormones (cortisol nor DHEAS), suggesting that the observed multiday fluctuations in stress hormones were not detectable by individuals. Indeed, if multiday hormonal fluctuations reflect an intrinsic homeostatic process, then it would be counterproductive for these oscillations to induce stress.

Hormonal dynamics are inherently oscillatory, and these dynamics are essential to healthy function^49^. The ultradian (pulsatile) and circadian rhythmicity of cortisol is well studied, and longer oscillations have been observed in timing and amplitude of cortisol secretion, including seasonal, menstrual and weekday changes^50,51^. While the purpose of longer, periodic fluctuations in hormone levels remains unclear, it has been hypothesised that they provide an adaptive advantage to respond to changing environmental demands, and similar rhythms have been observed in other biomarkers of healthy individuals, including blood pressure, basal temperature and immune activation^52^. The current results suggest that, while intrinsic, multiday fluctuations of cortisol were maintained in epilepsy, overall cortisol levels were higher and morning cortisol levels were aligned with seizure cycles, with peak seizure risk times corresponding to increased morning cortisol for 11 out of 12 people. Importantly, individuals with stronger seizure cycles exhibited a stronger association between seizure cycle phase and cortisol concentrations. Conversely, evening cortisol was not correlated with multiday seizure cycles. This difference in morning vs. evening samples appears to reflect the tendency for evening cortisol to be influenced by seizures occurring around the time of sampling, and potentially other acute stressors encountered during the day, compared to morning cortisol.

It is possible that seizure susceptibility is increased at times when cortisol is naturally elevated or, conversely, other factors that heighten seizure risk may also modulate cortisol. In the latter case, disruption of cortisol dynamics may cause further damage, as networks underlying hormonal secretion are highly sensitive to the timing of stressors^49^. For instance, perturbations of the HPA-axis and its coupled oscillators may induce longer-term disruptions to sleep, menstruation and fertility^49,53^. More generally, misalignment of hormonal rhythms is implicated in a range of diseases along with overall morbidity/mortality. Further work is needed to directly probe whether multiday fluctuations in cortisol drive or are co-modulated with seizure cycles.

A major limitation of the current study was the reliance on self-reported seizure timing to establish seizure cycles. Although cycles of seizure diaries can reliably correlate with cycles estimated from electrographic seizures^28^, it is likely that both under- and over-reporting of seizures affected the results. For instance, a more accurate record of seizure times would improve the model’s ability to detect seizure-related changes in cortisol. On the other hand, the use of self-reported seizures allows individuals with stronger cycles to be identified from routinely collected clinical data, potentially allowing targeted interventions aimed at mitigating stress for patients who may be most likely to benefit. Furthermore, while previous studies have shown acute post-ictal cortisol increase, the current study failed to observe significant changes related to seizures. There was a non-significant trend for increased evening cortisol around seizure times, which may become more pronounced with more accurate seizure capture. Similarly, allocated high/low risk periods were not associated with changes in cortisol, suggesting that prospective assignment of risk based on seizure diaries was not sensitive enough to track epilepsy nor autonomic imbalance.

Further work is underway to replicate the current study design in cohorts with chronic, implanted EEG recordings, to assess the relationship between multiday cycles of interictal discharges, seizures and stress. Furthermore, this study did not consider the effect of anti-seizure medications on cortisol, and individuals were on diverse treatments including monotherapy (4 individuals), polytherapy (7 individuals) or no medications (2 individuals). Previous studies have shown anti-seizure medications may affect hormone levels, as well as seizure cycles^28,54^, so further work is necessary to investigate these effects on the stress-seizure relationship. Similarly, the cohort was too small to robustly determine whether other demographic or clinical factors impact the findings relating stress and seizure cycles.

## Conclusion

The current study revealed important insights into the stress-seizure dynamic over slow timescales, providing the first line of evidence that stress hormones co-oscillate with multiday rhythms of seizure susceptibility, irrespective of perceived stress levels. This dissociation between perceived stress and physiological stress responses suggests multiday hormonal fluctuations reflect an intrinsic homeostatic process, which may be disrupted by external stressors like seizures. The results underscore the need for further longitudinal studies to shed light on the drivers of multiday physiological cycles in epilepsy, paving the way for more targeted interventions during periods of increased seizure susceptibility.

## Data Availability

All data produced in the present study are available upon reasonable request to the corresponding author.

## Authors’ Contributions

All authors read and approved the final version of the manuscript.

RES conceived of and led the study, contributed to data collection, data and statistical analyses, code generation, conception of ideas, writing, and editing of the text.

JNF conceived of and led the study, contributed to data collection, data analysis, funding acquisition, conception of ideas, writing, and editing of the text.

DBG contributed to conception of ideas, data analysis and editing of the text.

WJD contributed to conception of ideas, clinical analysis and editing of the text.

IG contributed to data analysis, statistical analysis and editing of the text.

DF contributed to conception of ideas, editing of the text and provided data collection resources.

EN contributed to editing of the text, clinical analysis and provided data collection resources.

MJC contributed to conception of ideas, funding acquisition, clinical analysis and editing of the text

PJK conceived of and led the study, contributed to data analysis, funding acquisition, conception of ideas, writing, and editing of the text.

## Ethics Committee Approval

Ethical approval for this study was granted by St Vincent’s Hospital Human Research Ethics Committee (HREC 009.19). All participants provided informed consent and were made aware that participation was voluntary, with the option to withdraw at any time.

