## Supplement for "Stress hormones are associated with multiday seizure cycles"

**Supplementary Table 1. 10-item Perceived Stress Scale scoring system.** Total score is the sum of scores for all responses.

| **Negative items (1, 2, 3, 6, 9, 10)** | | **Positive items (4, 5, 7, 8)** | |
| --- | --- | --- | --- |
| **Possible response** | **Score per response** | **Possible response** | **Score per response** |
| Never | 0 | Never | 4 |
| Almost never | 1 | Almost never | 3 |
| Sometimes | 2 | Sometimes | 2 |
| Fairly often | 3 | Fairly often | 1 |
| Very often | 4 | Very often | 0 |

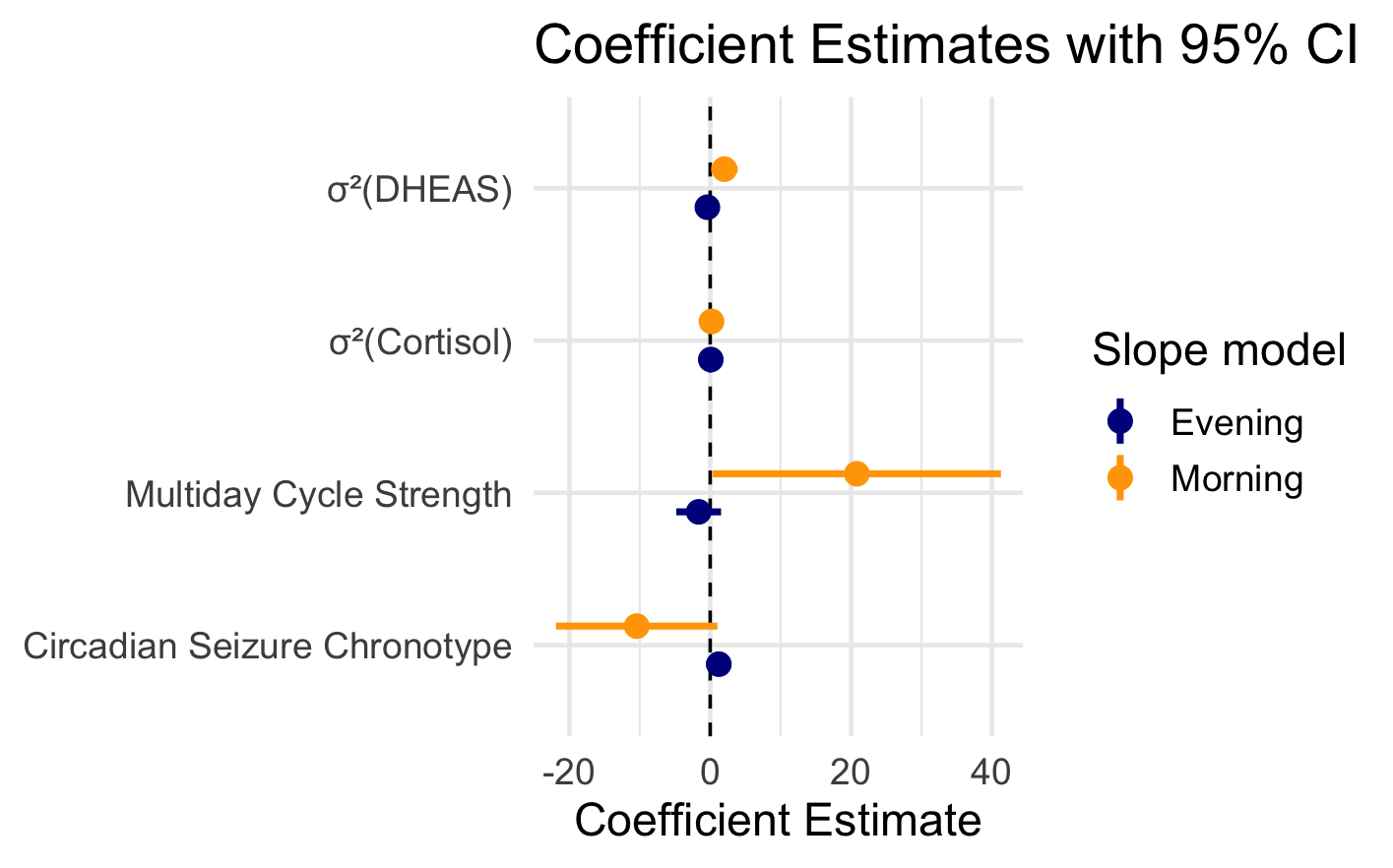

**Supplementary Figure 1.** Coefficient estimates and 95% confidence intervals (CIs) for fixed effects in the linear models predicting the slope of cortisol change across multiday seizure cycles. Two models were evaluated and plotted together: morning slopes (orange) and evening slopes (blue). Variance symbols (σ^2^) represent individuals’ variances in DHEAS and cortisol concentrations across all saliva samples; Multiday Cycle Strength represents the magnitude of the Synchronization Index; and Circadian Seizure Chronotype represents the tendency of individuals’ seizures to occur in the morning or afternoon, using the direction of the mean resultant vector (1=morning seizures, occurring between 12AM and 12PM; 0=afternoon seizures, occurring between 12PM and 12AM).

**Supplementary Table 2.** Coefficient estimates, confidence interval (CI) and p-values values for fixed effects in the linear models predicting the slope of cortisol change across multiday seizure cycles. Significant (p < 0.05) values are bolded. Variance symbols (σ^2^) represent intra-individual variances in DHEAS and cortisol concentrations across all saliva samples; Multiday Cycle Strength represents the magnitude of the Synchronization Index; and Circadian Seizure Chronotype represents the tendency of individuals’ seizures to occur in the morning or afternoon, using the direction of the mean resultant vector (1=morning seizures, occurring between 12AM and 12PM; 0=afternoon seizures, occurring between 12PM and 12AM).

|  |  | **Coefficient estimate** | **CI (low)** | **CI (high)** | **p-value** |
| --- | --- | --- | --- | --- | --- |
| **Morning cortisol slope model** | σ^2^(DHEAS) | **2.02** | **0.14** | **3.89** | **0.038** |
|  | σ^2^(Cortisol) | 0.18 | -0.02 | 0.38 | 0.067 |
|  | Multiday Cycle Strength | **20.82** | **0.36** | **41.28** | **0.047** |
|  | Circadian Seizure Chronotype | -10.44 | -21.89 | 1.02 | 0.068 |
| **Evening cortisol slope model** | σ^2^(DHEAS) | **0.09** | **0.05** | **0.12** | **<0.001** |
|  | σ^2^(Cortisol) | **-0.40** | **-0.69** | **-0.11** | **0.014** |
|  | Multiday Cycle Strength | -1.64 | -4.82 | 1.54 | 0.263 |
|  | Circadian Seizure Chronotype | 1.23 | -0.56 | 3.01 | 0.148 |

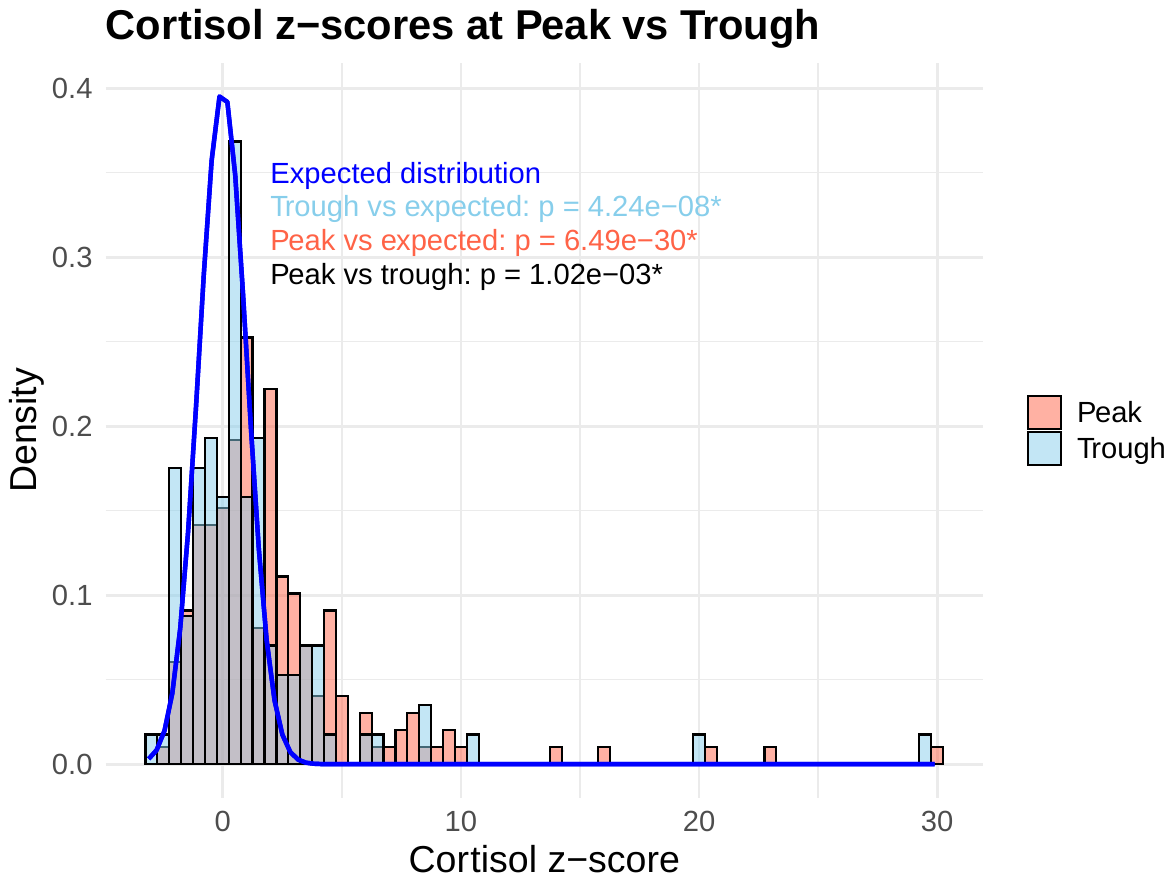

**Supplementary Figure 2. Cortisol levels in the epilepsy population at multiday seizure cycle peak vs. trough, compared to the expected general population.** The blue histogram of represents all participants’ z-scored cortisol concentrations taken from saliva samples collected during the trough of their multiday cycle, whereas the red histogram represents saliva samples collected during the peak of their multiday cycle. The dark blue “Expected distribution” line represents expected distribution of cortisol levels across the general population (provided by Nutripath Pty Ltd). Differences in the distributions were tested using the **Kolmogorov-Smirnov test and p-values (* indicating significance at p < 0.05) are reported.**

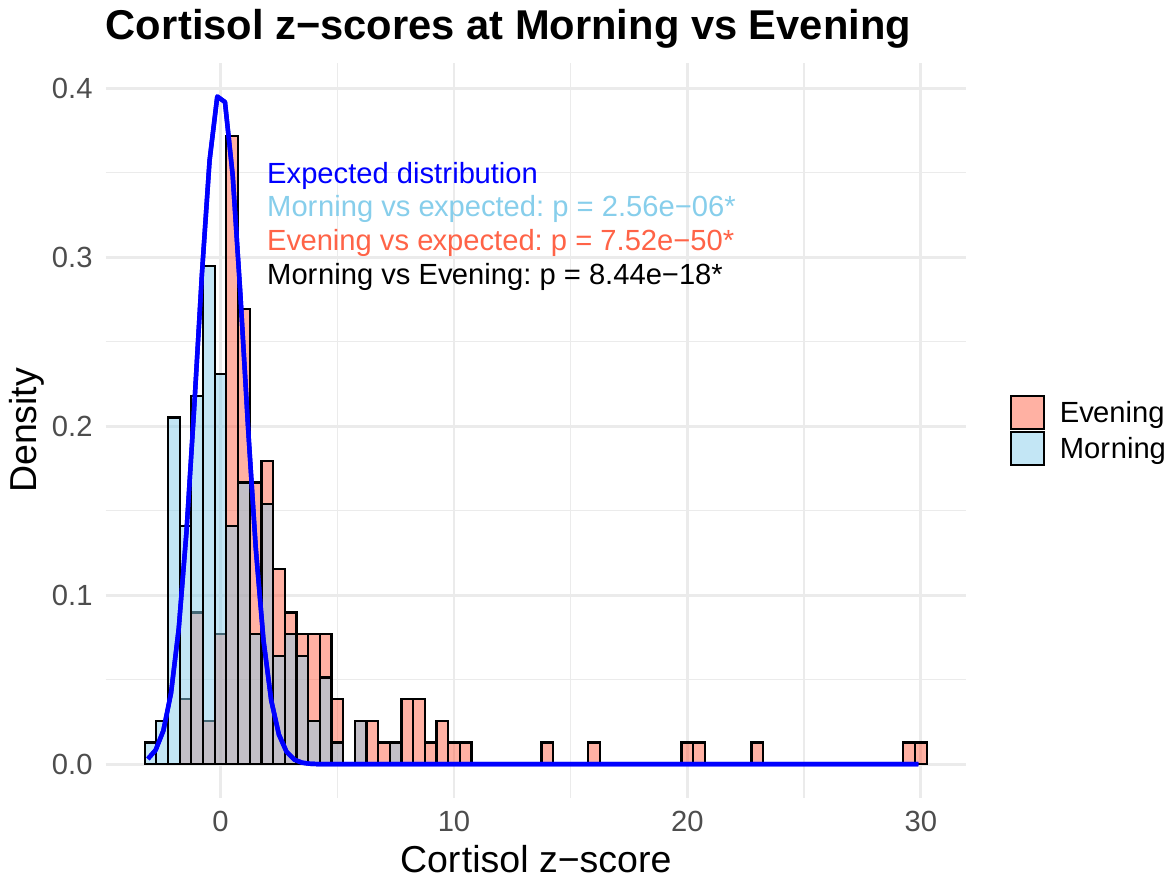

**Supplementary Figure 3. Cortisol levels in the epilepsy population in morning vs. evening samples, compared to the expected general population.** The blue histogram of represents all participants’ z-scored cortisol concentrations taken from saliva samples collected during the morning, whereas the red histogram represents saliva samples collected during the evening. The dark blue “Expected distribution” line represents expected distribution of cortisol levels across the general population (Nutripath Pty Ltd etc.). Differences in the distributions were tested using the **Kolmogorov-Smirnov test and p-values (* indicating significance at p < 0.05) are reported.**

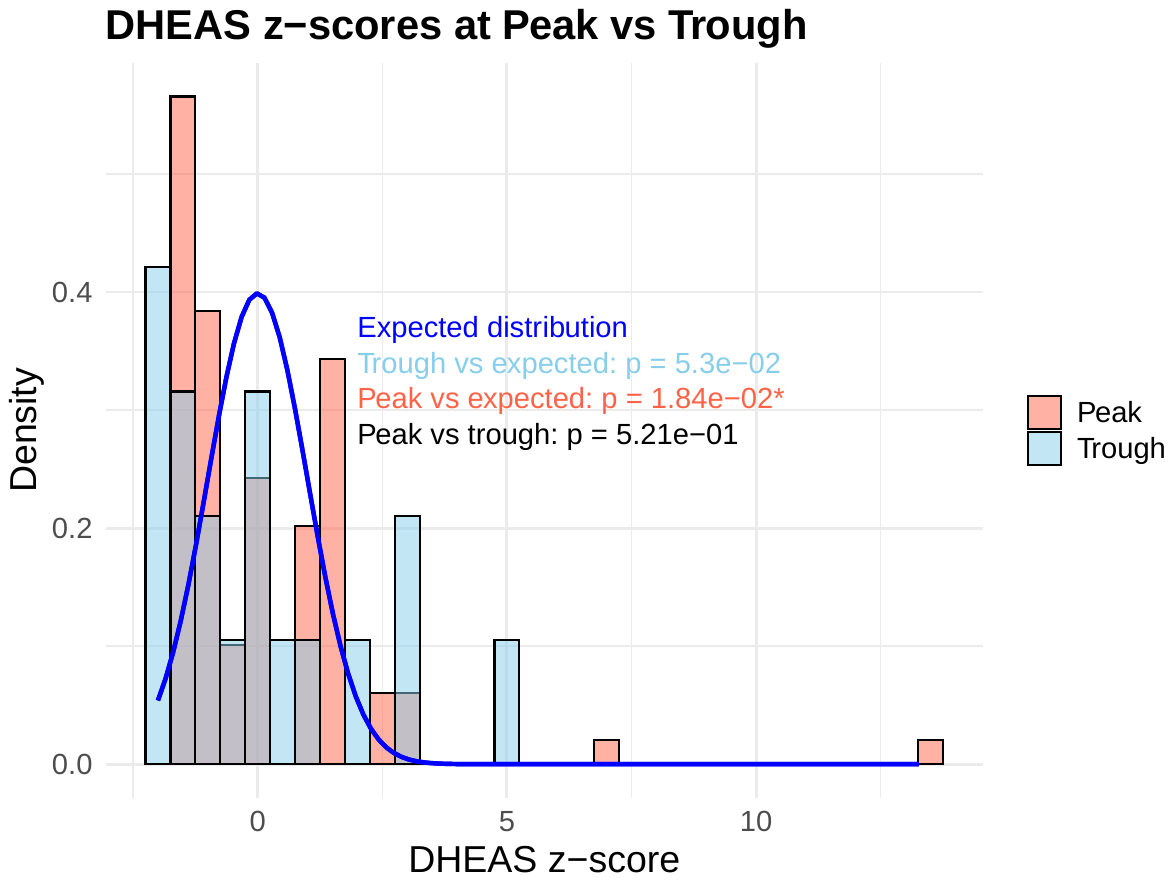

**Supplementary Figure 4. DHEAS levels in the epilepsy population at multiday seizure cycle peak vs trough, compared to the expected general population.** The blue histogram of represents all participants’ z-scored DHEAS concentrations collected during the trough of their multiday cycle, whereas the red histogram represents saliva samples collected during the peak of their multiday cycle. The dark blue “Expected distribution” line represents expected distribution of DHEAS levels across the general population (Nutripath Pty Ltd etc.). Differences in the distributions were tested using the **Kolmogorov-Smirnov test and p-values (* indicating significance at p < 0.05) are reported. Note that** DHEAS concentrations were only measured in the first saliva sample of each sampling collection period (i.e. Day 1, 8 AM), thus only 4 DHEAS saliva collections were obtained per person.

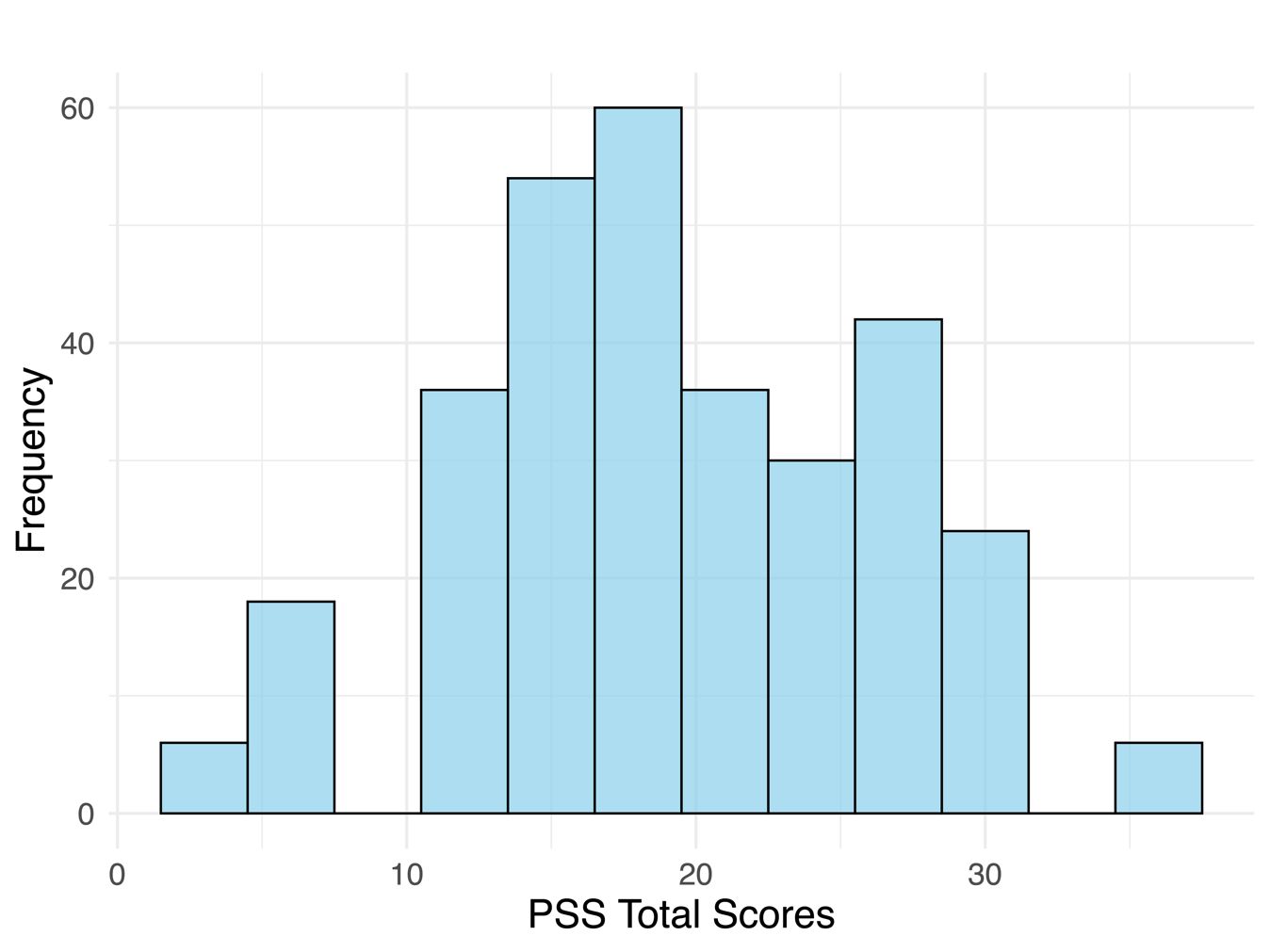
**Supplementary Figure 5.** Histogram of participants’ PSS scores across all time periods.

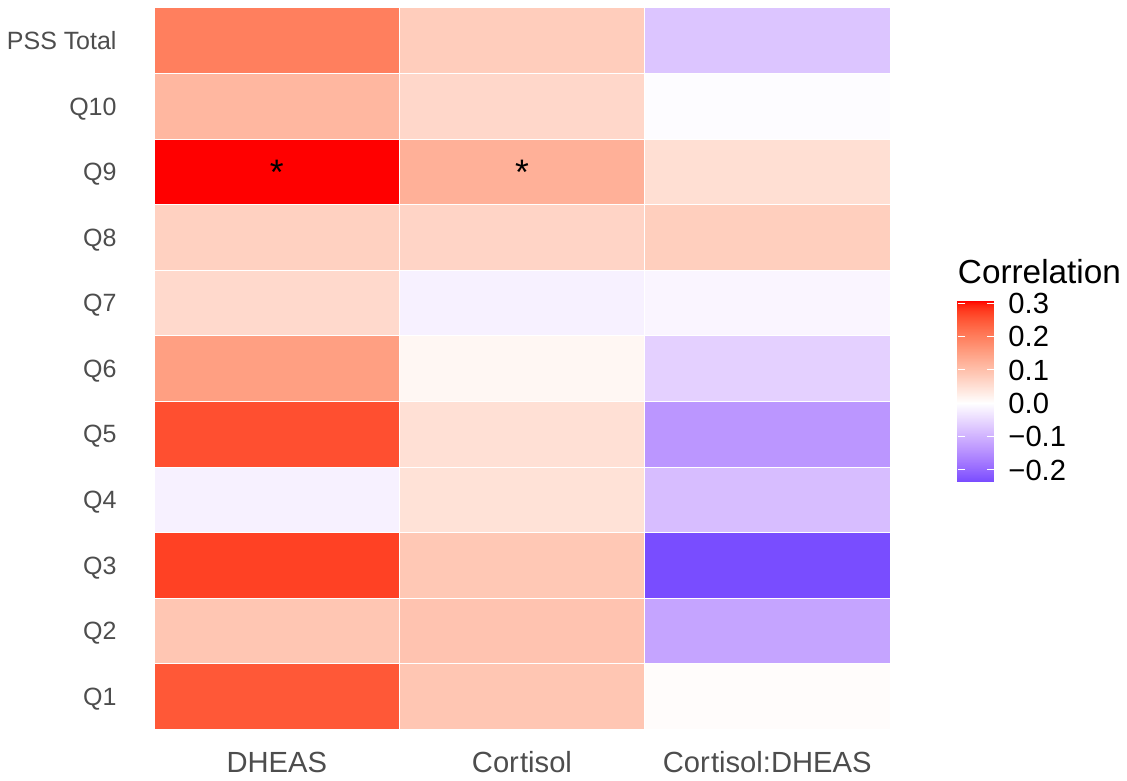

**Supplementary Figure 6. Correlation matrix to assess relationships between individual items on the Perceived Stress Scale (PSS) questionnaire and stress hormone concentrations (DHEAS, Cortisol and Cortisol:DHEAS ratio).** * Indicates significant correlation (p<0.05) using Pearson’s correlation coefficient.
